# Immune Cell Exhaustion and Apoptotic Markers in Major Depressive Disorder: Effects of in Vitro Cannabidiol Administration

**DOI:** 10.1101/2024.12.24.24319614

**Authors:** Muanpetch Rachayon, Ketsupar Jirakran, Pimpayao Sodsai, Chavit Tunvirachaisakul, Atapol Sughondhabirom, Jing Li, Yingqian Zhang, Michael Maes

**Affiliations:** Department of Psychiatry, Faculty of Medicine, Chulalongkorn University and King Chulalongkorn Memorial Hospital, the Thai Red Cross Society, Bangkok, Thailand; Center of Excellence for Maximizing Children’s Developmental Potential, Department of Pediatrics, Faculty of Medicine, Chulalongkorn University, Bangkok, Thailand; Center of Excellence in Immunology and Immune-Mediated Diseases, Department of Immunology, Faculty of Medicine, Chulalongkorn University and King Chulalongkorn Memorial Hospital, Bangkok, Thailand; Sichuan Provincial Center for Mental Health, Sichuan Provincial People’s Hospital, School of Medicine, University of Electronic Science and Technology of China, Chengdu 610072, China; Key Laboratory of Psychosomatic Medicine, Chinese Academy of Medical Sciences, Chengdu, 610072, China; Kyung Hee University, 26 Kyungheedae-ro, Dongdaemun-gu, Seoul 02447, Korea; Department of Psychiatry, Medical University of Plovdiv, Plovdiv, Bulgaria; Research Institute, Medical University Plovdiv, Plovdiv, Bulgaria; Research and Innovation Program for the Development of MU - PLOVDIV– (SRIPD-MUP), Creation of a network of research higher schools, National plan for recovery and sustainability, European Union – NextGenerationEU

## Abstract

**Background:** Immune dysregulation is a component of Major Depressive Disorder (MDD). Cannabidiol (CBD) has immunomodulatory and putative antidepressant effects. The relationship between apoptotic and exhaustion immune markers and the clinical features of MDD and the effects of CBD on these markers are still unknown.

**Objectives:** To assess pro-apoptotic (CD95) and T cell exhaustion (TIM-3) markers on immune cells in patients with MDD, as well as the impact of in vitro CBD administration on these markers.

**Methods:** We recruited healthy controls and MDD patients and evaluated the immunophenotypes of T/B lymphocytes using flow cytometry in unstimulated and anti-CD3/CD28 stimulated conditions. We evaluated the immune profiles of M1 macrophages, T helper (Th)17 cells, immune-inflammatory response system (IRS), T cell proliferation, and immune-related neurotoxicity (IRN). We investigated the in vitro effects of CBD on immune cell subsets at concentrations of 0.1 µg/mL, 1 µg/mL, and 10.0 µg/mL.

**Results:** The stimulated CD3+CD95+ cell percentages were substantially correlated with the number of depressive episodes, recurrence of illness, and suicidal behaviors. The stimulated CD8+TIM-3+ cell percentages were substantially and inversely associated with the M1, IRS, CIRS, T cell growth, and IRN immune profiles. TIM-3+ bearing CD3+, CD4+ and CD8+,cells were significantly suppressed by lower CBD concentrations (0.1 – 1 µg/mL). TIM-3+ and all CD95+ bearing cells, with the exception of CD3+CD95+, were suppressed by the higher CBD concentrations.

**Discussion:** Aberrations in immune checkpoint molecular processes impact the features of MDD. CBD significantly impacts apoptotic and exhaustion processes thereby possibly interfering with immune homeostasis.

## Introduction

Major depressive disorder (MDD) is among the most common psychiatric disorders globally. Recent studies indicate that MDD is associated with immunological dysregulation (1, 2). MDD is linked to the activation of the immune-inflammatory response system (IRS), as indicated by elevated levels of pro-inflammatory cytokines, including interleukin (IL)-1β, IL-6, interferon (IFN)-γ, and tumor necrosis factor (TNF)-α (3, 4). MDD is also associated with heightened activity of the compensatory immune-regulatory system (CIRS) (3). The latter may mitigate excessive inflammation by downregulating the IRS and generating anti-inflammatory cytokines such as IL-4, IL-10, and transforming growth factor (TGF)-β(3). In the acute phase of severe MDD, the IRS is more activated than the CIRS, leading to net activation of the IRS (3). A prior flow cytometry investigation corroborates this hypothesis, demonstrating that individuals with MDD exhibit heightened activation of T helper (CD4+) and cytotoxic T cells (CD8+), accompanied by a reduction in T regulatory (Treg) cells (5). Treg cells are a crucial element of the CIRS. The dysregulation of these systems during MDD episodes may result in heightened activation of M1 macrophages, T helper (Th)1 and Th17 cells, elevated concentrations of their neurotoxic cytokines, and an expanded population of CD4+CD40L+ T cells capable of inducing neurotoxicity (3, 5). Moreover, increased numbers of activated T cells and lowered Treg cell numbers are significantly associated with key features of MDD, namely the number of depressive episodes and suicidal attempts, lifetime suicidal behaviors (that is a composite based on suicidal ideation and attempts), recurrence of illness (ROI, that is a composite based on the number of episodes, suicidal behavior and ideation), and the severity of the phenome of the index episode (a composite based on severity of depression, anxiety and suicidal behaviors) (6).

The activation of the IRS is governed not only by the CIRS pathways, involving Th2 and Treg cells, but also by apoptotic events and T cell exhaustion (7). Two key pathways that govern apoptosis and the exhaustion of immune cells are CD95 and TIM-3 (8). CD95, or the Fas/APO-1 receptor, is essential for initiating apoptosis and significantly influences IRS activity. Upon activation by its ligand, CD95 ligand (CD95L), CD95 forms the death-inducing signaling complex (DISC). This complex comprises CD95, FADD, procaspase-8, procaspase-10, and c-FLIP, all of which are crucial for the initiation of apoptosis (8, 9). CD95-mediated apoptosis is essential for immune surveillance, facilitating the removal of aberrant cells and averting illnesses such as cancer and autoimmune disorders (10). Patients with treatment-resistant depression exhibit a reduced fraction of CD3+CD8+CD95+ T cells, suggesting a possible imbalance in apoptotic pathways and immunological function (11–13).

TIM-3, or T cell immunoglobulin mucin-3, is an immune-regulatory protein which expression is elevated on exhausted T cells (14). Furthermore, TIM-3 modulates apoptosis by interacting with its receptor, galectin-9, resulting in calcium influx into Th1 cells, which initiates apoptosis and suppresses Th1-mediated immune responses (15, 16). A study by Qin et al. indicates that TIM-3 is associated with chronic stress, promoting autophagy, and leading to stress-induced immunosuppression (17). Nevertheless, the associations between CD95 and TIM- 3 bearing immune cells and the features of MDD, including suicidal behaviors, recurrence of illness (staging), severity of the index episode, have remained unknown.

Recent research indicates that cannabidiol (CBD), a phytocannabinoid derived from the cannabis plant, may possess antidepressant characteristics, as evidenced in animal models of depression (18, 19). CBD functions via the endocannabinoid system, which comprises cannabinoid receptors, chiefly cannabinoid receptor 1 (CB1) and cannabinoid receptor 2 (CB2) (20). CB1 receptors are predominantly located in the central nervous system, with lower levels in peripheral blood cells. In contrast, CB2 receptors are primarily located on peripheral immune cells, such as B lymphocytes (21). The regulation of CB1 and CB2 receptors is recognized to affect IRS functions (22–24). CB1 receptors, despite their lower prevalence in T cells, are crucial for T cell growth and death (23). Both CB1 receptor agonists and antagonists demonstrate anti-inflammatory effects, highlighting CB1’s dual function in immune regulation (25, 26). Exogenous CB1 agonists are recognized for their immunosuppressive and anti-inflammatory properties, although CB1 antagonists also have anti-inflammatory effects, highlighting the intricacy of CB1-mediated immune responses (23, 25, 26). CB2 receptors are present on peripheral blood mononuclear cells (PBMC) (27, 28), and exogenous CB2 receptor agonists exert anti-inflammatory effects via multiple pathways, such as lowering pro-inflammatory cytokines, decreasing reactive oxygen species, attenuating T cell activation, and slowing microglial migration (29–32). Previous studies have also identified a dose-dependent effect of CBD, with higher doses exhibiting a more obvious immunosuppressive effect through CB1/CB2 receptors (33, 34). However, there is no research on CBD’s effects on the number of immune cells bearing CD95+ and TIM-3+ markers.

Hence, this study is performed to examine a) the relationship between T cell apoptosis and exhaustion cell markers and MDD; and b) the effects of in vitro CBD administration on the number of T cells bearing CD95+ and TIM-3+.

## Methodology

### Participants

The research included subjects from Chulalongkorn University in Bangkok. Senior psychiatrists from the Department of Psychiatry enlisted outpatients diagnosed with MDD, while healthy controls were recruited via posters and referrals. All participants filled out questionnaires and submitted blood samples. The inclusion criteria for patients comprised persons aged 18 to 65 who comprehended Thai, had received a diagnosis of MDD from a psychiatrist utilizing DSM-5 criteria, and possessed a Hamilton Depression Rating Scale (HAMD) score exceeding 17, signifying moderate to severe depression. Patients were excluded if they had other psychiatric disorders such as schizophrenia, schizoaffective disorder, obsessive-compulsive disorder, post-traumatic stress disorder, psycho-organic disorders, or substance use disorders. Healthy controls were matched for age, sex and educational attainment and were not included if they had any axis 1 DSM-IV-TR condition or a familial history of MDD, BD, or psychosis. Patients and controls were excluded if they exhibited allergic or inflammatory responses in the preceding three months, had neuroinflammatory, neurodegenerative, or neurological disorders (e.g., epilepsy, Alzheimer’s disease, multiple sclerosis, Parkinson’s disease), (auto)immune diseases (e.g., COVID-19 infection, chronic obstructive pulmonary disease, inflammatory bowel disease, psoriasis, type 1 diabetes, asthma, rheumatoid arthritis), a history of immunomodulatory drug usage, therapeutic doses of antioxidants or omega-3 supplements within three months, or anti-inflammatory drug usage within one month prior to the study. Women who are pregnant or nursing were also excluded. A subset of MDD patients were administered psychotropic drugs, comprising sertraline (18 patients), various other antidepressants (8 patients; fluoxetine, venlafaxine, escitalopram, bupropion, mirtazapine), benzodiazepines (22 patients), atypical antipsychotics (14 patients), and mood stabilizers (4 patients). The statistical analyses were adjusted for any effects of the drug state.

All participants provided written informed consent, and the study was conducted in accordance with international and Thai ethical and privacy legislation. The Institutional Review Board of the Faculty of Medicine at Chulalongkorn University sanctioned the study (#528/63), in compliance with the Declaration of Helsinki, the Belmont Report, CIOMS Guidelines, and the International Conference on Harmonization in Good Clinical Practice (ICH-GCP).

## Measurements

### Clinical assessments

A psychiatrist performed semi-structured interviews. This interview included demographic data such as age, gender, body mass index, number of education years, smoking behaviors, and the incidence of depressive episodes. The diagnosis of Major Depressive Disorder (MDD) was established utilizing the DSM-5 criteria (35). Psychiatric illnesses were identified in each patient utilizing the Mini-International Neuropsychiatric Interview (M.I.N.I.) (36). The same physician administered the 17-item Hamilton Depression Rating Scale (HAM-D) to assess the severity of depressive symptoms (37). The Thai version of the State–Trait Anxiety Inventory (STAI) was utilized to assess the intensity of state anxiety. The Columbia-Suicide Severity Rating Scale (C-SSRS) lifeline version was employed to evaluate the degree of current and lifetime suicidal ideation among participants, as well as any history of suicide attempts (38).

According to Maes et al. (6), lifetime suicidal behaviors were calculated as the first principal component derived from lifetime C-SSRS items concerning suicidal ideation (SI) and suicidal attempts (SA). A composite score was generated utilizing both assessments of SI and SA, termed lifetime suicidal behaviors (SB). ROI was determined by the first principal component derived from the quantity of depressive episodes, the C-SSRS item about "lifetime suicidal ideation," and the C-SSRS item concerning the total number of actual lifetime suicide attempts. The severity of the phenome of MDD was calculated as outlined by Maes et al. (6). To do this, we utilized the latent variable scores derived from the first principal component retrieved from the HAMD and STAI scores, along with current suicidal behaviors. The latter scores were calculated using 9 C-SSRS questions that evaluated recent (past month) reports of suicidal ideation and attempts. Tobacco use disorder was diagnosed based on DSM-5 criteria. Weight (in kilograms) was divided by height (in square meters) to get body mass index (BMI).

### Methodology

Blood samples were collected from participants after an overnight fast, between 8:00 and 9:00 a.m. using BD Vacutainer® EDTA (10 mL) and BD Vacutainer® SST™ (5 mL) tubes from BD Biosciences (Franklin Lakes, NJ, USA). Serum was obtained by allowing the serum-separating tubes to clot at room temperature for 30 minutes, followed by centrifugation at 1,100 g for 10 minutes at 4°C. PBMCs were isolated from the blood samples using density gradient centrifugation for 30 minutes at 1,500 rpm with Ficoll® Paque Plus (GE Healthcare Life Sciences, Pittsburgh, PA, USA). Cell counts and viability were determined with a hemocytometer and trypan blue (0.4% solution, pH 7.2-7.3, Sigma-Aldrich Corporation, St. Louis, Missouri, United States), ensuring over 95% cell viability under all conditions. For PBMC activation, 96-well plates were prepared by coating them overnight with 5 µg/mL of anti-human CD3 antibody (OKT3, eBioscience). Each well was then seeded with 3 x 10^5^ PBMCs and 5 μg/mL of anti-human CD28 antibody (CD28.2, eBioscience). The cells were cultured in RPMI 1640 medium with L-glutamine, supplemented with 10% fetal bovine serum (FBS) and 1% penicillin-streptomycin (Gibco Life Technologies, Carlsbad, CA, USA), for 3 days at 37°C in a 5% CO_2_ incubator. Unstimulated PBMCs cultured under the same conditions were used as a negative control. After the 3-day culture period, lymphocyte immunophenotypes were determined using flow cytometry. The effects of CBD were determined with 3 concentrations: 0.1 µg/ml, 1 µg/mL, and 10 µg/mL. We assessed the percentage of 6 different phenotypes, namely CD3+CD95+, CD3+Tim3+, CD4+CD95+, CD4+Tim3+, CD8+CD95+, CD8+Tim3+cells and this in 5 conditions: unstimulated or baseline, anti-CD3 and anti-CD28 stimulated, stimulated + 0.1 µg/mL CBD, stimulated + 1.0 µg/mL CBD and stimulated + 10.0 µg/mL CBD. To study lymphocyte immunophenotypes, cultured PBMCs were labeled with monoclonal antibodies, including CD3-PEcy7, CD4-APCcy7, CD8-APC, CD95-PE/Dazzle594 and Tim3-PE (Biolegend, BD Biosciences and R&D Systems). We stained cells with all antibodies above which are surface markers for 30 minutes at 4°C in the dark. Cells were washed 3 times by 1X PBS supplemented with 2% FBS. The flow cytometry was performed using LSRII flow cytometer (BD Biosciences) to evaluate lymphocyte immunophenotypes. All data were analyzed using FlowJo X software (Tree Star Inc., Ashland, OR, USA). Electronic Supplementary File (ESF), Figure 1 shows our gating strategies.

As previously described (39), we measured cytokines/chemokines/growth factors in unstimulated and stimulated diluted whole blood culture supernatant using the same blood samples as those employed for the flow cytometry. We used RPMI-1640 medium supplemented with L-glutamine, phenol red, and 1% penicillin (Gibco Life Technologies, USA), with or without 5 µg/mL PHA (Merck, Germany) + 25 µg/mL lipopolysaccharide (LPS; Merck, Germany). On sterile 24-well plates, 1.8 mL of each medium was combined with 0.2 mL of 1/10- diluted whole blood. Using the Bio-Plex Pro human cytokine 27-plex assay kit (BioRad, Carlsbad, California, United States of America) and the LUMINEX 200 instrument (BioRad, Carlsbad, California, United States of America), the cytokines/chemokines/growth factors were measured. The intra-assay CV values were less than 11%.

## Statistics

Analysis of variance (ANOVA) was utilized to evaluate continuous variables across categories, whilst chi-square tests were conducted to ascertain associations among categorical variables. The various impacts of time or groups on immune profiles underwent a false discovery rate (FDR) p correction (40). Multiple regression analysis was employed to identify the relationships between the features of depression (dependent variables) and the unstimulated and stimulated immune cell types, while allowing for the effects of age, sex, BMI, smoking and the drug state of the patients. To achieve this, we employed a manual and an automated method with a p-entry threshold of 0.05 and a p-removal threshold of 0.06 while evaluating the changes in R^2^. Multicollinearity was assessed using tolerance and variance inflation factor, whilst multivariate normality was evaluated by Cook’s distance and leverage, and homoscedasticity was examined via the White and modified Breusch-Pagan tests. The results of the regression analysis were consistently bootstrapped using 5,000 bootstrap samples, and these are given if the findings were incongruent. We utilized generalized estimating equations (GEE) analysis with repeated measurements to assess the effect of CBD administration on the immune cell subsets. The predetermined GEE analyses, employing repeated measures (unstructured correlation matrix, linear scale response, and maximum likelihood estimation for the scale parameter), incorporated fixed categorical effects of time (unstimulated, stimulated, and stimulated with the three CBD concentrations), diagnostic groups (depression versus controls), time-by-group interaction, sex, smoking, and continuous fixed covariates, specifically age and BMI. The repeated measurements of immune cell subtypes were the dependent variables. We calculated the estimated marginal mean (SE) values for the treatment and diagnostic groups, as well as the treatment x group interactions, and employed protected pairwise contrasts (least significant difference at p=0.05) to assess differences between the treatment conditions and the interaction patterns between group and treatment. Additionally, we incorporated the drug state into the GEE analysis as supplementary predictors to eliminate any influence of these potential confounders. The GEE technique allows for the consideration of significant confounders in the analysis of treatment effects at the individual level. There were no missing values in any of the clinical, immune, or sociodemographic data. The tests were two-tailed, with statistical significance established at p <0.05. The statistical studies were conducted using IBM SPSS version 28 for Windows.

Employing a two-tailed test with a significance level of 0.05 and assuming a power of 0.80, an effect size of 0.2, two groups, and five measurements with intercorrelations of around 0.4, the estimated sample size for a repeated measures ANOVA would be approximately 38.

## Results

### Demographic and clinical data

**Table 1** displays the sociodemographic and clinical information of the patients and the control group in the study. The two groups exhibited no significant differences in age, gender distribution, educational attainment, or smoking behaviors. Nevertheless, patients exhibited a somewhat elevated BMI in comparison to the controls. We controlled all findings for potential confounding variables, including age, BMI, sex, smoking, and medication status. However, no significant impacts from these potential confounding factors were detected. The patients had markedly elevated average HAMD and STAI scores compared to the controls, signifying that the majority of patients were undergoing moderate to severe clinical depression and anxiety. All clinical characteristic ratings for depression were considerably elevated in MDD compared to controls.

**Table 1.** Demographic and clinical data of the depressed patients and healthy controls (HC) included in the present study.

| Variables | HC (n=19) | MDD (n=29) | F/X<sup>2</sup> | df | P |
| --- | --- | --- | --- | --- | --- |
| Sex (Male/Female) | 6/13 | 11/18 | 0.20 | 1 | 0.653 |
| Age (years) | 33.8 (8.2) | 28.9 (8.6) | F=3.94 | 1/46 | 0.053 |
| Education (years) | 16.0 (2.3) | 15.7 (1.4) | F=0.53 | 1/46 | 0.473 |
| Smoking (Yes/No) | 17/2 | 23/6 | FEPT | - | 0.366 |
| BMI (kg/m<sup>2</sup>) | 21.5 (2.5) | 25.0 (6.0) | F=6.19 | 1/46 | 0.017 |
| HAMD | 1.0 (1.6) | 23.4 (5.8) | MWUT | 1/46 | <0.001 |
| STAI | 37.7 (10.9) | 56.7 (7.3) | 51.83 | 1/46 | <0.001 |
| Number of depressive episodes | 0 | 2.03 (0.94) | MWUT | - | <0.001 |
| Number of suicidal attempts | 0 | 1.24 (1.55) | MWUT | - | <0.001 |
| Lifetime suicidal behaviors | -0.987 (0.0) | 0.624 (0.758) | MWUT | - | <0.001 |
| Recurrence of illness index | -1.089 (0.0) | 0.731 (0.560) | MWUT | - | <0.001 |
| Total phenome score | -1.131 (0.338) | 0.747 (0.408) | 277.14 | 1/46 | <0.001 |

Results are shown as mean ( ± SD). FEPT: Fisher's exact probability test; X<sup>2</sup>: analysis of contingency tables, F: results of analysis of variance, MWUT: Mann-Whitney U test; BMI: body mass index; HAMD: Hamilton Depression Rating Scale score.

### Baseline and stimulated lymphocyte population in MDD

ESF, Table 1 shows that no notable alterations were detected in other immune profiles, including CD3+, CD4+, and CD8+ cells expressing CD95+ or TIM3+, in both baseline and stimulated circumstances.

### Associations between MDD features and number of CD95 and TIM-3 bearing cells

**Table 2** presents the outcomes of multiple regression analysis with clinical features as dependent variables. We identified substantial correlations between the proportion of CD3+CD95+ cells under stimulated circumstances and several clinical characteristics. The quantity of depressive episodes had a significant negative correlation with CD3+CD95+%, which explained 9.2% of the variance. A comparable trend was noted with the frequency of suicide attempts, exhibiting a significant negative impact that accounted for 10.1% of the variance. CD3+CD95+% accounted for 15.5% of the variance in lifetime suicidal behaviors, demonstrating an inverse correlation. A higher percentage of CD3+CD95+ cells was strongly correlated with a lower ROI. The severity of the phenome exhibited a notable negative correlation with the CD3+CD95+ T cells, accounting for 10.5% of the variance.

**Table 2.**
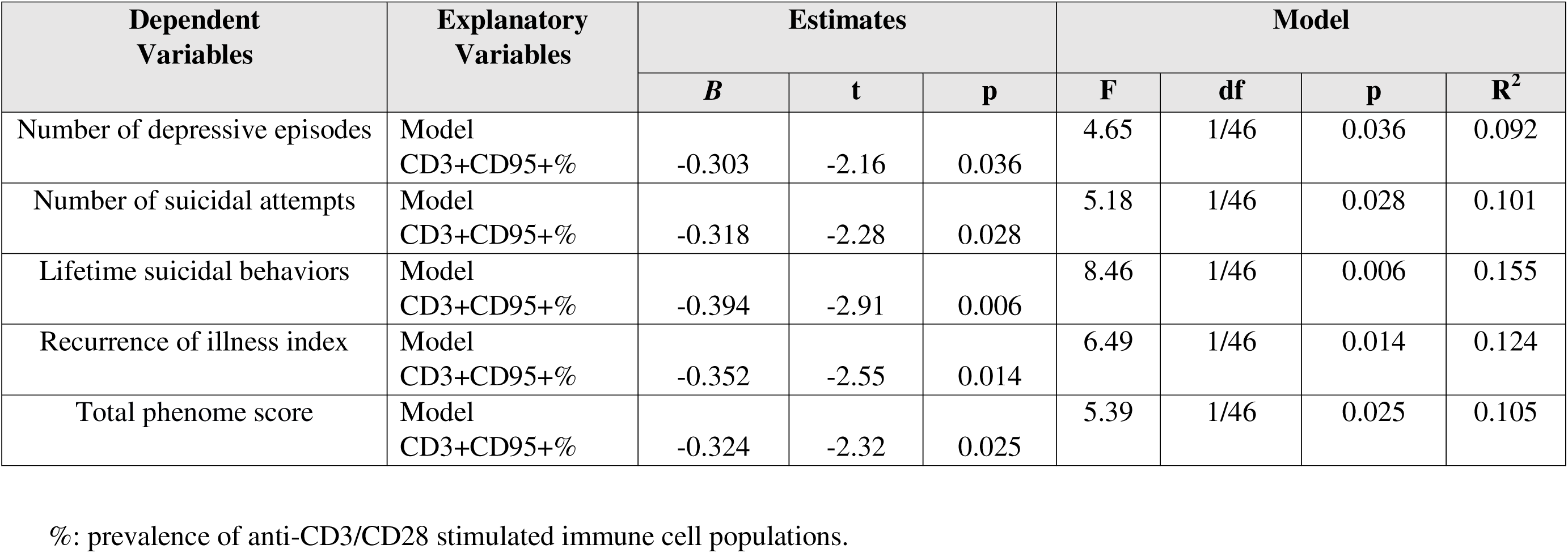
Results of multiple regression analyses with the clinical features of depression as dependent variables.

| Dependent Variables | Explanatory Variables | Estimates |  |  | Model |  |  |  |
| --- | --- | --- | --- | --- | --- | --- | --- | --- |
|  |  | *B* | *t* | *p* | *F* | *df* | *p* | *R*<sup>2</sup> |
| Number of depressive episodes | Model<br>CD3+CD95+% | -0.303 | -2.16 | 0.036 | 4.65 | 1/46 | 0.036 | 0.092 |
| Number of suicidal attempts | Model<br>CD3+CD95+% | -0.318 | -2.28 | 0.028 | 5.18 | 1/46 | 0.028 | 0.101 |
| Lifetime suicidal behaviors | Model<br>CD3+CD95+% | -0.394 | -2.91 | 0.006 | 8.46 | 1/46 | 0.006 | 0.155 |
| Recurrence of illness index | Model<br>CD3+CD95+% | -0.352 | -2.55 | 0.014 | 6.49 | 1/46 | 0.014 | 0.124 |
| Total phenome score | Model<br>CD3+CD95+% | -0.324 | -2.32 | 0.025 | 5.39 | 1/46 | 0.025 | 0.105 |

%; prevalence of anti-CD3/CD28 stimulated immune cell populations.

Overall, the proportion of CD3+CD95+ cells under stimulated conditions consistently acted as a substantial negative predictor in all models, underscoring its potential impact on the severity of clinical features of MDD.

### Regression analyses of immune profiles on numbers of CD95+ and TIM-3+ cells

We conducted multiple regression analysis with immunological profiles (M1, Th-1, Th- 17, IRS, T cell growth, and neurotoxicity) as dependent variables and the percentages of CD95+/TIM-3+ lymphocytes as the explanatory variable under both stimulated (S) and unstimulated (U) circumstances. **Table 3** indicates that, under stimulated conditions, the fraction of CD8+TIM3+ cells adversely affected the immunological profiles across the different models. First, elevated percentages of activated CD8+TIM3+ cells correlated with diminished expression of cytokine profiles, including M1, IRS, and CIRS, accounting for 8.2%, 9.1%, and 8.9% of the variation, respectively. The activated percentage of CD8+TIM3+ cells had a negative correlation with T cell proliferation and immune-related neurotoxicity (IRN), explaining 8.5% and 13.4% of the variation, respectively.

**Table 3.**
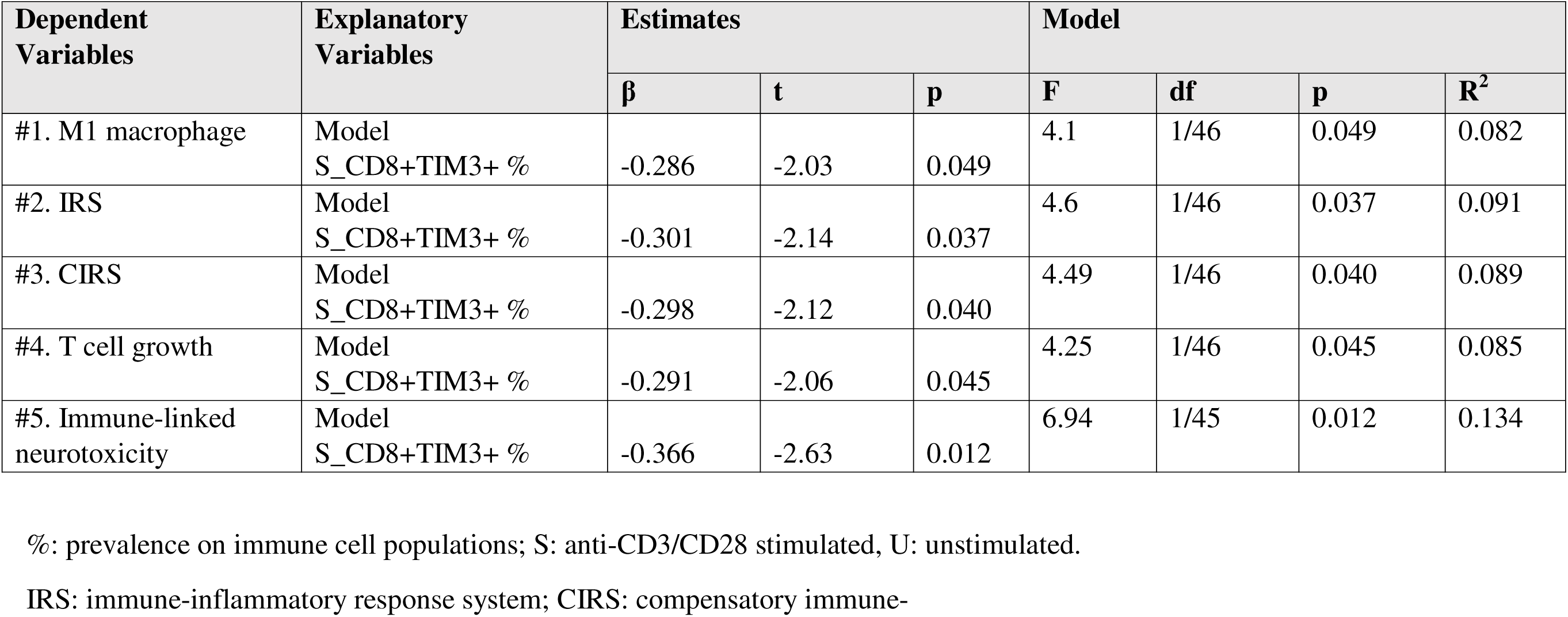
Results of multiple regression analyses with immune profiles as dependent variables and immune cell expression markers as explanatory variables.

### Effects of cannabidiol on lymphocyte and pro-apoptotic markers percentages

**Table 4** illustrates the impact of CBD on the proportions of CD95+ and TIM3+ T cells. GEE analysis was employed to assess all subjects across five experimental conditions: unstimulated, stimulated with anti-CD3/anti-CD28 antibodies, and incubated with three varying doses of CBD. The findings underscore the impact of CBD on these immune cell subgroups. At lower dosages (0.1 µg/mL and 1.0 µg/mL), CBD selectively inhibited TIM3-expressing cells, encompassing the CD3+TIM3+, CD4+TIM3+, and CD8+TIM3+ subsets. Moreover, at these concentrations, only the CD8+CD95+ fraction exhibited suppression. At a concentration of 10 µg/mL, CBD exhibited significant suppressive effects on TIM3+ and CD95+ expressing CD3+, CD4+, and CD8+ cells. GEE analysis showed that the overall stimulated number of CD3+CD95+ cells was significantly (p=0.041) lower in MDD (estimated marginal mean: 69.50 ±1.28) than in controls (74.33 ±1.82).

**Table 4.** Effects of cannabidiol (CBD) on anti-CD3/CD-28 induced changes in various CD95+ or TIM3+ bearing immune cells.

| Variables | Unstimulated | Stimulated (St)<br>□ | St + CBD<br>0.1 µg/mL □ | St + CBD<br>1.0 µg/mL □ | St + CBD<br>µg/mL 10 □ | Wald x2<br>(df=4) | p<br>Value |
| --- | --- | --- | --- | --- | --- | --- | --- |
| CD3+CD95+% | 13.93 (2.96) | 93.11 (0.56)<sup>d</sup> | 92.83 (0.61)<sup>d</sup> | 92.71 (0.78)<sup>d</sup> | 67.02 (2.49) □ □<br>□ | 361.54 | <0.001 |
| CD3+TIM3+% | 4.25 (1.09) | 61.03 (1.17)<sup>b c d</sup> | 59.57 (1.27)<sup>a c</sup> □ | 57.24(1.30) □ □<br>□ | 15.40 (1.25) □ □<br>□ | 1501.52 | <0.001 |
| CD4+CD95+% | 18.24 (3.21) | 94.30 (0.54) □ | 93.95 (0.67) □ | 94.13 (0.91) □ | 69.47 (2.64) □ □<br>□ | 331.85 | <0.001 |
| CD4+TIM3+% | 2.95 (1.04) | 54.91 (1.30)<sup>b c d</sup> | 53.36 (1.45)<sup>a c</sup> □ | 50.89(1.50)<sup>a</sup> □ □ | 11.33 (1.01) □ □<br>□ | 1468.82 | <0.001 |
| CD8+CD95+% | 8.93 (2.94) | 98.59 (0.18)<sup>b</sup> □ | 98.83 (0.17)<sup>a</sup> □ □ | 98.66 (0.23)<sup>b</sup> □ | 92.31 (1.41) □ □<br>□ | 334.23 | <0.001 |
| CD8+TIM3+% | 5.23 (1.66) | 77.63 (1.38)<sup>b</sup> □ □ | 76.28 (1.49)<sup>a</sup> □ □ | 73.63 (1.60) □ □<br>□ | 20.96 (1.34) □ □<br>□ | 828.95 | <0.001 |

All results of GEE analysis examining the effects of time (unstimulated versus stimulated) and cannabidiol (CBD) effects on immune cell subsets bearing CD95+ and TIM+. Results are shown as estimated marginal mean values (SE). All unstimulated immune cell numbers are significantly lower than the 4 stimulated values. <sup>a,b,c,d</sup>: protected post-hoc differences among the 4 stimulated values (at p=0.05).

## Discussion

### Associations between pro-apoptotic markers and MDD

This study’s first major finding is the negative correlation between CD95 expression on CD3+ T cells under stimulated conditions and several clinical outcomes in depression, including ROI, total phenome score, frequency of depressive episodes, and lifetime suicidal behaviors. A reduced expression of CD95 was associated with elevated scores in these outcomes, indicating that CD95 may have protective effects attenuating depressive symptoms and reduce the likelihood of relapse and suicidal behaviors.

Limited research has investigated the correlation between MDD and CD95 expression. A study revealed that patients with treatment-resistant depression demonstrated a reduced proportion of CD3+CD8+CD95+ cells (41). A study by Szuster-Ciesielska et al. investigated apoptosis levels in the blood leukocytes of individuals with depression, revealing an increase in apoptosis correlated with heightened expression of CD95 (42). Similarly, CD95 dysregulation is apparent in other immune-related illnesses that demonstrate considerable comorbidity with MDD. Genetic mutations in CD95 or CD95L are linked to systemic lupus erythematosus (SLE)

(10). A significant decrease in Fas receptor expression has been seen in multiple sclerosis (MS) patients, with receptor levels rising following treatment, indicating that T cells in MS displayed reduced sensitivity to apoptosis prior to intervention (43).

CD95 functions as a death receptor that triggers apoptosis upon interaction with CD95L, an essential mechanism for maintaining immune tolerance and homeostasis (44, 45). Research by Paulsen et al. demonstrates that increased CD95L levels facilitate death, whilst reduced levels encourage T cell activation (46). Thus, abnormal operation of the CD95/CD95L complex may lead to immunological dysregulations observed in several diseases (47, 48). Previously, we observed that patients with MDD demonstrated heightened T cell activation and dysregulation of Treg cells (5). The cumulative effects of compromised Treg functions and dysfunctional apoptotic mechanisms together with the heightened T cell activation in MDD may exacerbate the pronounced IRS activation in that illness.

### TIM-3, immunoregulation and MDD

The second major finding is the robust negative connection between the number of TIM- 3 bearing CD8+ cells and cytokine production, encompassing that of M1 macrophages, IRS, CIRS, Th17, T cell proliferation, and IRN immune cell profiles.

It is known that TIM-3 attenuates T-cell receptor signaling in CD8+ T cells by blocking nuclear-factor κB (49). In addition, TIM-3 serves as a hallmark of T cell exhaustion and operates as a co-inhibitory immune checkpoint protein which attenuates IRS and autoimmune responses, regulates macrophage functions, and promotes immunological tolerance (50–52). In rheumatoid arthritis, TIM-3 has been identified as having a negative connection with disease severity, indicating its involvement in the regulation of Treg cells to promote immunotolerance (53). Furthermore, in MS patients, the impaired negative regulation of TIM-3 and diminished expression relative to healthy individuals was restored following treatment (54). Numerous investigations suggest that TIM-3 exerts an inhibitory role, especially in chronic inflammatory situations (50, 51). Given the negative relationship between the immunological profiles assessed in the current study and TIM-3, it can be posited that a diminished inhibitory action via this exhaustion mechanism may result in enhanced activation of immune cells.

Consequently, the cumulative effects of T cell activation, and compromised Treg functions coupled with dysfunctional apoptotic processes in MDD may be further aggravated by loss of TIM-3 checkpoint regulation.

### CBD aggravates inflammation in a concentration-dependent manner

The third major finding of the current study is that CBD doses ranging from 0.1 to 1.0 µg/mL exhibited an inhibitory effect on TIM3+ expressing CD3+, CD4+, and CD8+ cells. In addition, at a dosage of 10 µg/mL, CBD demonstrated a stronger inhibitory impact on CD3+/CD4+/CD8+CD95+ cells. Although these findings correspond with CBD’s established immunomodulatory characteristics, they contrast with its anticipated anti-inflammatory effects. Numerous studies indicate that impairing CD95 activity can lead to immunological dysregulation (55, 56). Consequently, elevated levels of CBD inhibit CD95 expression on lymphocytes, potentially diminishing apoptotic checkpoint control thereby disturbing immunological homeostasis and fostering inflammation (46). Furthermore, at lower dosages (0.1–1.0 µg/mL), CBD exhibited an inhibitory effect on TIM-3 expression in immune cells, and this effect was more pronounced at the higher concentrations. Consequently, CBD administration at therapeutical dosages could further exacerbate T cell activation and inflammation by downregulating TIM-3 (34). A prior investigation into the impact of CBD on cytokines demonstrated that CBD exerts an immunological modulatory effect in a concentration-dependent way on several cytokines, with concentrations of 1.0 µg/mL or greater potentially inhibiting the production of CIRS activity coupled with increased growth factor production, while higher doses suppressed Th1, Th17, IRS, and CIRS cytokine profiles, but elevated growth factor and T cell growth profiles (34).

Endogenous cannabinoids, including anandamide (AEA) and 2-arachidonoylglycerol (2- AG), are recognized for their role in regulating inflammation and cellular death (57, 58). AEA demonstrates considerable affinity for both CB1 and CB2 receptors, functioning as a partial agonist with greater affinity for CB1 compared to CB2. Conversely, 2-AG functions as a complete agonist at both CB1 and CB2 receptors, but with moderate to low affinity for both receptors (59, 60). While AEA is acknowledged for its anti-inflammatory capabilities (57, 61), the involvement of 2-AG in inflammation is more complex and may also show pro-inflammatory effects (62, 63).

Our work suggests that CBD may increase immunological activation in depression by interfering with the apoptotic and exhaustion processes.

Limitations.

Our findings would have been even more interesting if we had assessed additional apoptosis-related markers or programmed death markers, including PD-1 (programmed death-1). Moreover, subsequent studies ought to examine these death, apoptosis and checkpoint markers at the partial remission and full remission stages. The intracellular processes elucidating the effects of CBD on the CD95+ and TIM-3+ cellular markers have to be clarified.

## Conclusions

The stimulated percentage of CD3+CD95+ cells is strongly and inversely correlated with essential characteristics of MDD, whether considering lifetime aspects (ROI, lifetime suicidal behaviors) or evaluations during the acute phase of MDD (severity of the phenome). The stimulated percentage of CD8+TIM3+ cells is negatively correlated with the activation of M1, IRS, CIRS, T cell proliferation, and IRN immunological profiles. These findings may indicate that reduced levels of checkpoint (apoptotic and exhaustion) markers in MDD may have diminished the negative feedback on immune activation pathways. Consequently, disruptions in apoptotic mechanisms in immune cells greatly influence the phenotype of MDD, whereas abnormalities in immunological exhaustion indicators may lead to diminished immunoregulatory effects. Consequently, CD95 and TIM-3 may represent novel pharmacological targets for the treatment of the acute phase of MDD.

Administering lower concentrations of CBD, specifically between 0.1 and 1.0 µg/mL, significantly suppressed TIM-3+ expressing CD3+, CD4+, and CD8+cells. Elevated doses of CBD had inhibitory effects on both TIM-3+ and CD95+ bearing cells. This suggests that CBD significantly affects indicators related to apoptosis and exhaustion. These inhibitory actions may diminish the negative feedback of the apoptosis and exhaustion checkpoint proteins on immunological activation in MDD, hence contributing to the pathophysiology of the acute phase of this disorder. Therefore, it cannot be recommended to use CBD as an anti-inflammatory compound.

## Author’s contributions

All the contributing authors have participated in the manuscript. M.M. and M.R. designed the study. M.R. and K.J. recruited patients and controls. P.S. performed assays A.S. secured financial support. M.R. wrote the first draft of this paper which was further worked out by M.M. K.J., P.S., C.T., A.S., J.L. and Y.Z. further edited the paper. All authors participated in the data interpretation and consented to the publication of the final paper version.

## Funding Statement

AMERI-ASIA MED CO, Ltd, supported this work.

## Ethical statement

All participants provided their written informed consent before their involvement in the study. The research was performed in compliance with the Declaration of Helsinki of 1975, as revised in 2013, and received approval from the Ethics Committee of the Institutional Review Board of the Faculty of Medicine, Chulalongkorn University, Bangkok, Thailand (#528/63).

## Data Availability Statement

The dataset produced and/or examined in this study will be accessible from the corresponding author (M.M.) upon reasonable request, following the complete utilization of the dataset by the authors.

## Conflicts of Interest

The authors declare no conflicts of interest with any commercial or other affiliations related to the submitted article.

## Supporting information

ESF

## Data Availability

All data produced in the present work are contained in the manuscript

---

**ESF, Figure 1.**
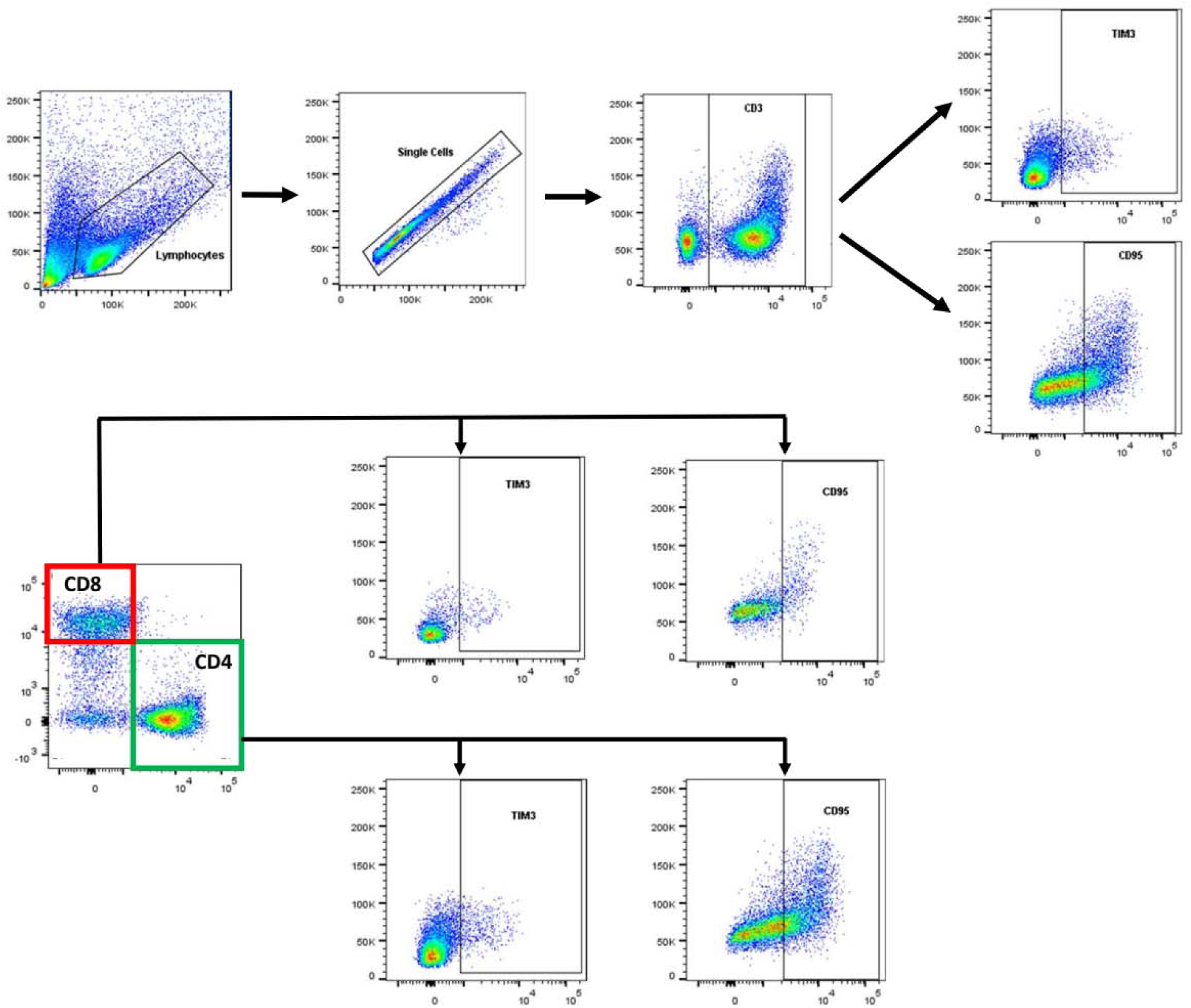
The gating strategy used in the current study.

**ESF, Table 1.** Differences in unstimulated (UNST) and stimulated (STIM) changes in the percentage of lymphocyte populations and death cells markers in healthy controls (HC) and major depressed patients (MDD)

| Variables | Condition | Diagnosis |  | Tests of Model effects |  |  |
| --- | --- | --- | --- | --- | --- | --- |
|  |  | HC <sup>a</sup> n=19 | MDD <sup>c</sup> n=29 | Effects | Wald (df=1) | p |
| CD3+CD95+% | UNST | 17.36 (5.39) | 10.58 (3.06) | G | 0.195 | 0.659 |
|  | STIM | 94.77 (1.12) | 91.51 (0.80) | GXT | 0.022 | 0.882 |
| CD3+TIM3+% | UNST | 4.63 (1.63) | 3.70 (1.40) | G | 0.190 | 0.663 |
|  | STIM | 61.13 (1.74) | 60.76 (1.54) | GXT | 1.045 | 0.307 |
| CD4+CD95+% | UNST | 21.11 (5.86) | 15.48 (3.29) | G | 0.010 | 0.921 |
|  | STIM | 95.70 (1.19) | 93.01 (0.71) | GXT | 0.037 | 0.847 |
| CD4+TIM3+% | UNST | 4.14 (1.62) | 1.80 (0.78) | G | 0.820 | 0.363 |
|  | STIM | 53.65 (1.94) | 56.20 (1.56) | GXT | 2.201 | 0.138 |
| CD8+CD95+% | UNST | 12.18 (5.32) | 5.71 (3.08) | G | 0.594 | 0.441 |
|  | STIM | 99.20 (0.57) | 98.01 (0.42) | GXT | 0.432 | 0.511 |
| CD8+TIM3+% | UNST | 5.25 (2.31) | 5.52 (2.24) | G | 2.740 | 0.098 |
|  | STIM | 79.89 (1.81) | 75.69 (1.67) | GXT | 1.271 | 0.260 |

## Notes

### Competing Interest Statement

The authors have declared no competing interest.

### Funding Statement

This study was funded by AMERI-ASIA MED CO, Ltd,

### Author Declarations

Ethics committee/IRB of Chulalongkorn University gave ethical approval for this work

