## Supplementary material for "Immune Cell Exhaustion and Apoptotic Markers in Major Depressive Disorder: Effects of in Vitro Cannabidiol Administration": ESF

**Electronic Supplementary File (ESF)**

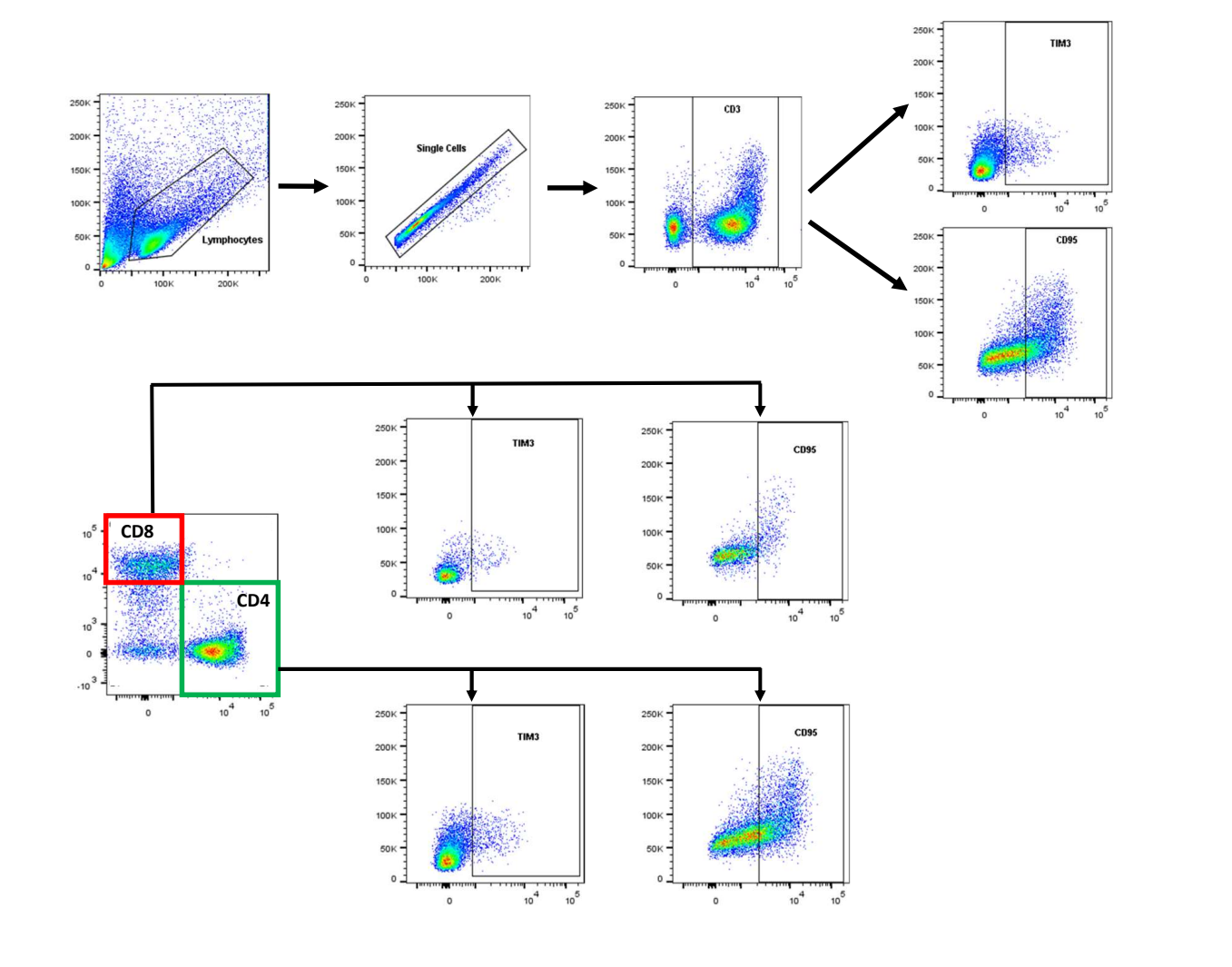

**ESF, Figure 1.** The gating strategy used in the current study.

**ESF, Table 1.** Differences in unstimulated (UNST) and stimulated (STIM) changes in the percentage of lymphocyte populations and death cells markers in healthy controls (HC) and major depressed patients (MDD)

|  | **Condition** | **Diagnosis** | | **Tests of Model effects** | | |
| --- | --- | --- | --- | --- | --- | --- |
| **Variables** |  | **HC ^a^ n=19** | **MDD ^c^ n=29** | **Effects** | **Wald (df=1)** | **p** |
| CD3+CD95+% | UNST | 17.36 (5.39) | 10.58 (3.06) | G | 0.195  0.022 | 0.659  0.882 |
|  | STIM | 94.77 (1.12) | 91.51 (0.80) | GXT |  |  |
| CD3+TIM3+% | UNST | 4.63 (1.63) | 3.70 (1.40) | G | 0.190  1.045 | 0.663  0.307 |
|  | STIM | 61.13 (1.74) | 60.76 (1.54) | GXT |  |  |
| CD4+CD95+% | UNST | 21.11 (5.86) | 15.48 (3.29) | G | 0.010  0.037 | 0.921  0.847 |
|  | STIM | 95.70 (1.19) | 93.01 (0.71) | GXT |  |  |
| CD4+TIM3+% | UNST | 4.14 (1.62) | 1.80 (0.78) | G | 0.820  2.201 | 0.363  0.138 |
|  | STIM | 53.65 (1.94) | 56.20 (1.56) | GXT |  |  |
| CD8+CD95+% | UNST | 12.18 (5.32) | 5.71 (3.08) | G | 0.594  0.432 | 0.441  0.511 |
|  | STIM | 99.20 (0.57) | 98.01 (0.42) | GXT |  |  |
| CD8+TIM3+% | UNST | 5.25 (2.31) | 5.52 (2.24) | G | 2.740  1.271 | 0.098  0.260 |
|  | STIM | 79.89 (1.81) | 75.69 (1.67) | GXT |  |  |
